# Effect of tetrabenazine on Stroop interference in Huntington’s disease: Pilot Open Label Trial

**DOI:** 10.1101/2025.07.09.25330323

**Authors:** Robert Fekete

## Abstract

**Objective:** Tetrabenazine has been shown to improve gating of abnormal visual stimuli and improve postural stability in Huntington’s disease (HD) patients as measured by computerized dynamic posturography testing. This open label pilot study aims to elucidate whether partial dopaminergic depletion via low dose tetrabenazine has a similar effect on masking out of abnormal visual stimuli on the Stroop interference test in HD patients. The study endpoint is the Golden formula calculated from off and on tetrabenazine Stroop test raw scores.

**Results:** Full data set was available from one subject with Stroop interference score -1.97 off tetrabenazine and -1 on tetrabenazine, with lower scores showing more interference. The data was not amenable to statistical analysis. A proposed statistical analysis is presented which may be useful to researchers seeking to utilize the methodology.

## Introduction

Tetrabenazine has been shown to improve gating of abnormal visual stimuli and improve postural stability in Huntington’s disease (HD) patients as measured by computerized dynamic posturography testing. This open label pilot study aims to elucidate whether partial dopaminergic depletion via low dose tetrabenazine has a similar effect on masking out of abnormal visual stimuli on the Stroop interference test in HD patients.

The effect of tetrabenazine on chorea has been well established [2]. Additional effects which may not necessarily involve chorea and may involve improvement in information processing of divergent stimuli have only recently been elucidated [1].

Showing an improvement in processing of visual stimuli at a lower dose would be an intriguing possibility, further supporting a separate, chorea-independent mechanism for some of tetrabenazine’s beneficial effects. Most likely, this is a dopamine related effect involving information processing in the frontal lobe [1]. Dopaminergic blockade in healthy volunteers with haloperidol has been shown to reduce Stroop interference [3]. Dietary dopaminergic depletion in healthy volunteers also reduced Stroop interference [4].

Analysis of raw scores from Stroop testing in the Huntington Study Group trial [2] showed a statistically significant decline in the raw word reading score, but no significant changes in the raw color reading or raw interference scores. Calculated Stroop scores using either the Golden or Chafetz formulae [5] were not reported for this trial. It may be possible that the calculated score would properly account for slower word reading in tetrabenazine treated Huntington disease patients and show reduction in interference even if the raw interference score remains unchanged. For this reason, raw as well as calculated interference scores via will be reported in this trial. A three-day withdrawal of steady state high tetrabenazine by Fekete et al. showed a significant effect on the raw Stroop test [6].

## Methods

The study was registered at the National Institutes of Health Protocol Registration and Results system as NCT01834911 on April 15^**th**^, 2013. The study was approved by the New York Medical College Office of Research Administration Institutional Review Board under tracking number L-10,830. There was no funding support for the trial. The recruitment period was between 11/12/2012 and 6/3/2017. Logistical issues delayed study registration.

Subjects who are residents of a nursing home and have an established diagnosis of Huntington disease were recruited by the principal investigator for participation in this study. Written informed consent was obtained from the subjects and also from next of kin given cognitive disturbance in HD.

Key inclusion criteria for the study were:

1. Established diagnosis of Huntington disease by movement disorders expert
2. Patients not currently taking tetrabenazine as well as those off tetrabenazine for at least three days
3. Patients should not have taken dopamine receptor blocking medication for at least three days (as this has been shown to affect the Stroop score in normals [3]).

The study was performed as a single arm medication de novo or withdrawal trial. Informed consent to participate was obtained from all participants and the next of kin. The data collection instrument is the standard Stroop color naming, word reading, and interference test.

Stroop test was administered to patients in the morning to patients who are off tetrabenazine (de novo or three-day withdrawal). 12.5 mg of TBZ will be administered just after the first Stroop test and also 3 hours later. On Stroop test was to be administered 6 hours after the off Stroop test.

The raw Stroop word reading, color reading, and interference scores were recorded and used for further analysis.

### Proposed Statistical analysis

The three raw Stroop scores will be compared between ON and OFF groups. In addition, both Golden interference scores will be calculated, with customary age correction according to established convention. The ON and OFF scores may be compared using paired Student’s t-test in the cases of raw scores and Golden calculated scores

Given that a study examining effect on Stroop scores in the setting of low dose tetrabenazine administration has not been previously performed, the size of the potential effect is not known and hence direct power analysis is not possible.

The closest study from which to obtain effect size on Stroop test for power analysis is a three day withdrawal of steady state tetrabenazine [6]. Using Glass’s delta formula:

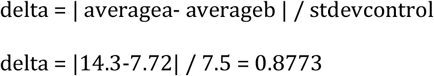

alpha=0.05 for single tailed t test power=1-beta error=0.95 with sample size of 8 as calculated by G Power software [7].

## Results

Two patients were enrolled in the study. Study was closed on 6/3/2017 given no further enrollment of subjects.

Full data set was available from one subject with Stroop interference score -1.97 off tetrabenazine and -1 on tetrabenazine, with lower scores showing more interference (Table 1). This subject was a 50 to 55 year old female with established diagnosis of HD by movement disorders expert who was not taking tetrabenazine. This subject’s score did not require age correction. There were no adverse events related to the trial.

**Table 1.** Results of raw Stroop Color, Word, Color Word, predicted Color Word (calculated as Color x Word / Color + Word), interference (Color Word – predicted Color Word), and t scores.

|  | Color | Word | Color Word | Predicted Color Word | Interference Score<br>Lower score worse indicating more interference | T score (50 average, normal range 40-60) | Percentile rank (Lower rank worse) |
| --- | --- | --- | --- | --- | --- | --- | --- |
| Patient 1 off medication | 14 | 25 | 7 | 8.97 | -1.97 | 48.03 | 42 |
| Patient 1 on medication | 6 | 3 | 1 | 2 | -1 | 47.8 | 41.3 |

## Limitations

This was an open label study. The study adheres to CONSORT Pilot Trial guidelines except that given rarity of this disorder and lack of funding, study resources supported only single arm medication de novo or withdrawal trial, without randomization. The data was not amenable to statistical analysis which severely limits the generalizability of study results. A proposed statistical analysis is presented which may be useful to researchers seeking to utilize the methodology. Future research can be performed in a double-blind fashion with a larger study cohort.

## Data Availability

All relevant data are within the manuscript and its Supporting Information files.

## Abbreviations

HD: Huntington’s disease (HD)
TCC: Terence Cardinal Cooke
TBZ: tetrabenazine

## Declarations

### Ethics Approval and Consent to Participate

The study adheres to CONSORT Pilot Trial guidelines except that given the rarity of this disorder and lack of funding, study resources supported only single arm medication withdrawal trial, without randomization and results were not amenable to statistical analysis. Completed CONSORT Pilot Trial checklist is available as Supplemental File 1.

The study is in compliance with the Declaration of Helsinki.

Written informed consent to participate was obtained from all participants and the next of kin. The study was registered at the National Institutes of Health Protocol Registration and Results system as NCT01834911. The study was approved by the New York Medical College Office of Research Administration Institutional Review Board under tracking number L-10,830.

## Consent for Publication

Not applicable

## Author Contributions

RF Conception, data acquisition, writing of manuscript.

## Data Availability

All data generated or analysed during this study are included in this published article.

## Acknowledgements

None

## Figure Legends

**Figure 1.**
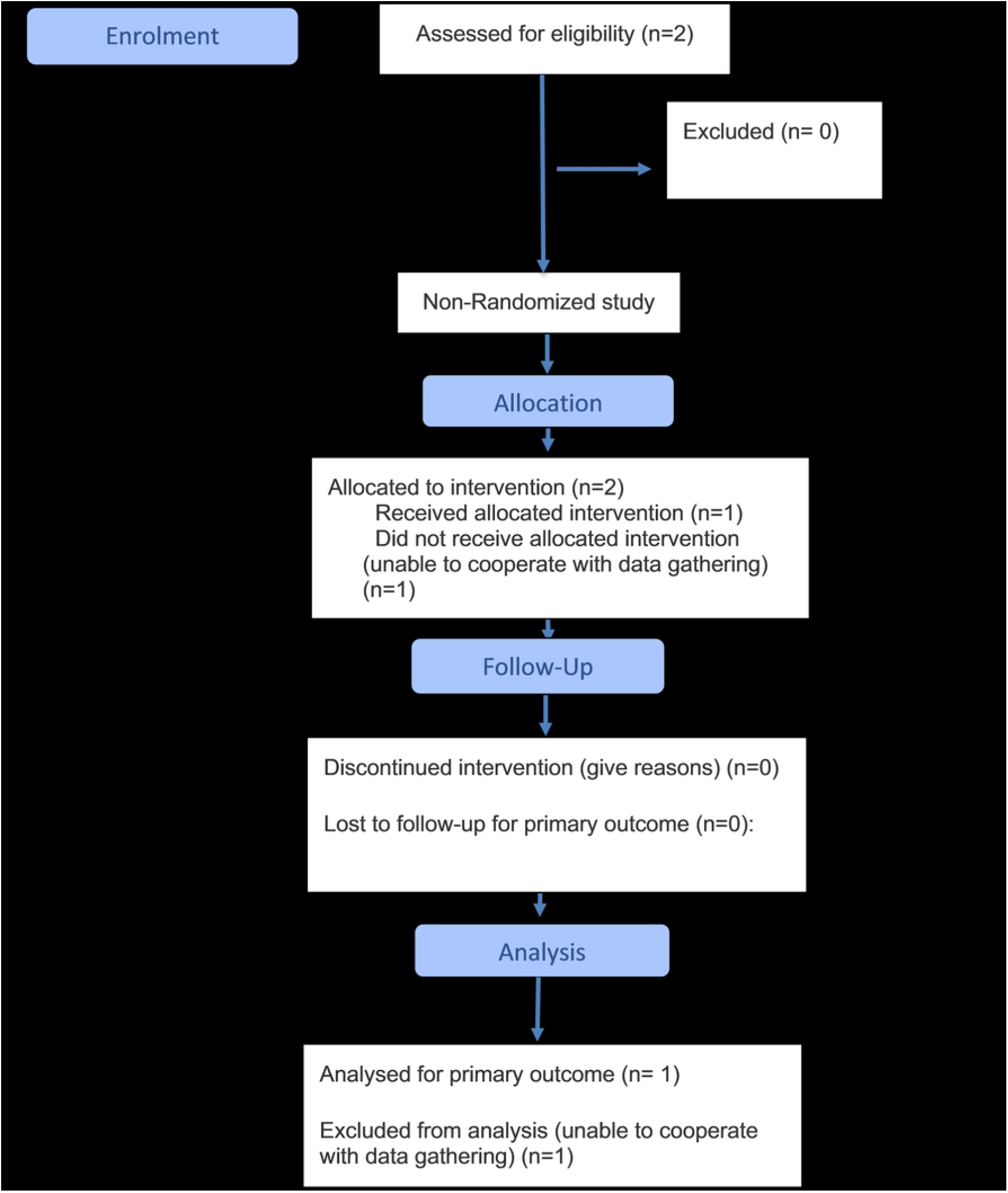
CONSORT Flow diagram [8].

## Supplemental Files

1. CONSORT Checklist

